# Modeling the Economic Impact of a CIZ1B Biomarker Blood Test for Lung Cancer Screening in High-Risk Populations

**DOI:** 10.1101/2025.01.28.25321291

**Authors:** Jennifer Hinkel, Rutuja Kavade, Madhura Joshi

**Affiliations:** Sigla Sciences

## Abstract

**Background:** This study evaluates the economic implications of incorporating a CIZ1B biomarker blood test into lung cancer screening protocols for Medicare-eligible high-risk populations. Despite the proven mortality reduction benefits of low-dose computed tomography (LDCT), its adoption remains low. Barriers such as access disparities, logistical challenges, and the high rate of false positives necessitating invasive follow-up limit LDCT’s broader acceptance. The addition of a blood-based biomarker test like CIZ1B could address these barriers by enhancing screening precision, reducing unnecessary diagnostic procedures, and increasing screening participation.

**Methods:** An economic model was developed to assess the cost savings and public health benefits of integrating CIZ1B in three scenarios: (1) sequential screening with LDCT following a positive biomarker test; (2) using CIZ1B to confirm LDCT-positive cases; and (3) an expanded screening paradigm where CIZ1B reduces barriers, increasing overall screening uptake. The model incorporates Medicare reimbursement rates, prevalence and screening sensitivity data, and cost estimates adjusted for inflation.

**Results:** Results indicate that integrating CIZ1B testing can yield net savings in the range of $500 Million by avoiding unnecessary biopsies and enabling earlier lung cancer detection and treatment. The expanded screening scenario projects additional savings through higher participation rates. This study highlights the potential of the CIZ1B biomarker to address critical challenges in lung cancer screening, reduce healthcare costs, and improve outcomes for high-risk populations.

## Introduction

Lung cancer screening with low-dose computed tomography (LDCT) for high-risk individuals is recommended by the US Preventative Services Task Force (USPST) and the National Comprehensive Cancer Network (NCCN), and is reimbursed by Medicare. (“Screening for Lung Cancer with Low Dose Computed Tomography (LDCT),” n.d.) Evidence indicates that regular screening of pre-symptomatic individuals in high-risk populations can achieve a significant reduction (20%) in mortality. (National Lung Screening Trial Research Team et al. 2011)

Despite established clinical benefits and cost-effectiveness, the implementation of screening programs faces substantial challenges, reflected in persistently low uptake rates across various healthcare settings. Studies in the US have shown uptake rates at only ∼4% to ∼6% of the eligible Medicare population. Low adoption rates are attributed to a variety of factors, including racial and socioeconomic disparities, geographic practice variation and geographic access barriers, inconvenience and availability of CT imaging, and anxiety around the frequency of false positives and subsequent invasive interventions.

The barriers to effective screening implementation for high-risk individuals are multifaceted and complex, despite Medicare coverage that results in zero out-of-pocket cost for the patient. The National Lung Screening Trial found that nearly a quarter of individuals require follow-up procedures to investigate suspicious imaging results, with approximately 96% of these investigations ultimately proving to be false positives. (National Lung Screening Trial Research Team et al. 2011)

This rate of false positives creates both personal and economic costs that make stand-alone imaging strategies unattractive to some patients and clinicians as a screening approach. (Goulart et al. 2012) The challenge is particularly acute among specific demographic groups, with documented disparities in screening rates among nonwhite Medicare beneficiaries and in certain geographic regions, notably the Southern United States. (Dwyer et al. 2024) A study led by City of Hope and Kaiser Permanente researchers in California examined barriers to LDCT for patients enrolled in a tobacco cessation program, and these included fear of test results, lack of awareness, fatalistic attitudes, radiation exposure fears, and smoking-related stigma. (Raz et al. 2019) Other studies cite logistical challenges, such as transportation difficulties, misunderstandings about insurance coverage, and perceived costs, present additional obstacles. (Jonnalagadda et al. 2012) (Carter-Harris et al. 2017).

Novel approaches to lung cancer screening, including blood-based biomarker tests, have the potential to mitigate identified barriers to LDCT screening while simultaneously offering health economic advantages. Additional screening methods may reduce expenditure on “unnecessary” biopsies among patients with false positive LDCT results and, if able to expand screening due to reduction of barriers, could identify more lung cancer cases at an early, more easily-treated stage. One of these blood-based biomarkers is Variant Ciz1 as described by Higgins et al. Variant Ciz1 is a strong candidate for a cancer-specific single marker capable of identifying early-stage lung cancer within at-risk groups without the need to perform invasive procedures (Higgins et al. 2012). Subsequent research on this biomarker has modeled estimated sensitivity and specificity for lung cancer and non-cancerous lung disease patients, which is pertinent to LDCT screening due to the false positive rate of identifying suspicious nodules and inflammation. (Coverley et al. 2017).

## Objective

This is a modeling study evaluating the economic and clinical impact of integrating CIZ1B biomarker testing into lung cancer screening protocols. The analysis focuses on Medicare beneficiaries eligible for lung cancer screening under current USPSTF and Medicare criteria, which as of 2022 include: Adults aged 50-77 years who are asymptomatic individuals (no signs or symptoms of lung cancer), current smokers or those who have quit within the past 15 years, a tobacco smoking history of at least 20 pack-years, and who receive an order for LDCT (“Screening for Lung Cancer with Low Dose Computed Tomography (LDCT),” n.d.).

The primary objective of this study is to model and evaluate the economic and public health impact of introducing a blood-based CIZ1B biomarker assay into the current lung cancer screening paradigm for high-risk individuals. Through comparative analysis of two implementation scenarios, we aim to:

1. Assess the potential cost savings of integrating CIZ1B testing with existing LDCT screening protocols among current Medicare beneficiaries receiving LDCT screening;
2. Model the potential impact of using CIZ1B as an initial screening tool that expands overall screening rates by reducing barriers to entry, with subsequent LDCT performed only in biomarker-positive cases; and
3. Quantify the potential reduction in unnecessary biopsies and associated cost savings with earlier detection of lung cancer across both scenarios.

The analysis specifically compares two baseline scenarios using sequential CIZ1B-LDCT testing at current screening rates (either before or after LDCT) against an expanded screening scenario where the availability of the blood-based biomarker test facilitates a 15% increase in initial screening rates prior to LDCT. This increase is modeled based on reduced barriers to entry and increased accessibility of blood-based testing compared to LDCT alone.

## Methods

This study built an economic model for cost savings in several implementation scenarios to assess potential real-world impact of a CIZ1B biomarker screening program in conjunction with LDCT for Medicare-eligible high-risk individuals.

### CIZ1B Biomarker Screening Implementation Scenarios

Scenario 1 modeled an initial CIZ1B biomarker test as a primary screen, followed by an LDCT for biomarker-positive cases only, and diagnostic follow-up costs for positive cases after LDCT including biopsy, at current LDCT screening rates among high-risk individuals (modeled at 4.6% of eligible individuals, as described below). Scenario 2 modeled the addition of CIZ1B testing as a sequential screening test for LDCT-positive cases at current LDCT screening rates among high-risk individuals.

Scenario 3 modeled an expanded testing scenario, with the same sequential protocol as Scenario 1, but projecting a modest 15% increase in screening uptake due to reduced barriers. Scenario 3 assumes that blood-based testing increases accessibility due to convenience, and that a sequential, layered screening approach that increases specificity further reduces prescriber hesitation to recommend screening, leading to increased uptake.

### Screening uptake

While the barriers to uptake are multi-factorial, we built a scenario that modeled a modest increase in screening program uptake (increase rates by 15% from 4.6% to 5.3%) due to the addition of the blood-based biomarker test. Our rationale for this includes the ability of a blood-based test to reduce geographic and logistic barriers such as availability of CT facilities or time and travel needed for screening, reduce the rate of false positives that lead to expensive and invasive diagnostic procedures, and reduce fear barriers around radiation exposure.

### Screening Uptake Rate Assumptions

We assumed a baseline screening uptake rate of 4.6% and an expanded screening rate of 5.3%. Studies in the past decade demonstrate a range of screening rates among eligible Medicare beneficiaries, depending on data sources, methods, and setting of care. In Medicare fee-for-service populations, a 2015-2016 analysis of claims showed a 3.3% uptake rate. (Tailor et al. 2020). A retrospective cohort study using national registry data from 2016 found a 4.1% screening rate. (Pham et al. 2020) National survey data from 2016-2017 demonstrated a 4.6% screening rate across the USA using the USPTF eligibility criteria. (Jemal and Fedewa 2017).

We selected the 4.6% rate because the authors of this paper felt that this best represented a national rate, understanding that claims data may show lower rates, but those studies are becoming less recent. We also acknowledge that studies in specific centers or health systems that have implemented programs to increase screening may show higher rates, but chose to model a conservative rate.

### Economic Modeling Approach

The economic model incorporated direct costs of screening and subsequent diagnostic follow-up to confirm or rule out a cancer diagnosis, as well as indirect costs such as savings derived from identifying and treating an early-stage lung cancer patient compared to diagnosing and treating that patient at a later stage of disease where more resource-intensive interventions are standard of care. Cost values were sourced from published literature as described in Table 1. Cost values from prior to 2020 were adjusted for inflation to 2024 dollars using the initial date as the latest date of data specified in the publication or, if the original publication adjusted for inflation to a more recent date, further adjustment was made from that later date. Outcomes modeled included the number of screening tests and biopsies conducted in each scenario, “unnecessary” biopsies avoided (biopsy procedures conducted on patients who did not have cancer), cost savings from reduced procedures, net cost impact in an expanded screening uptake scenario, and cost savings due to increased early diagnosis in an expanded screening uptake scenario. Table 2 describes additional assumptions and values selected as model inputs based on the literature.

**Table 1:** Cost Assumptions and Sources.

| **Component** | **Cost Assumption** | **Source** |
| --- | --- | --- |
| CIZ1B Biomarker test | \$295 | Provided by Cizzle Bio (manufacturer) |
| LDCT Screening | \$241 | Medicare physician fee schedule |
| Diagnostic follow-up to confirm or rule-out lung cancer | \$8,162 | (Lokhandwala et al. 2017)<br>With adjustment for inflation 2011 to 2024 |
| Estimated cost of lung cancer treatment per year, Stage I diagnosis | \$20,235 | (Mukherjee et al. 2022)<br>Weighted average calculated across surgery, radiotherapy, and ablation interventions |
| Estimated cost of lung cancer treatment per year, Stage IV diagnosis | \$129,977 | (Beata Korytowsky et al. 2018) |

**Table 2:** Population, Prevalence, Sensitivity, and Specificity Inputs and Assumptions.

| Component | Assumption & Rationale | Source |
| --- | --- | --- |
| Number of eligible Medicare beneficiaries | 4,900,000 (as of 2014) — this is a conservative estimate as the total Medicare population has grown significantly since 2014 | (Pyenson et al. 2014) |
| Prevalence of lung cancer in the screened Medicare population | 1.5% | (Pinsky et al. 2014) |
| Sensitivity of CIZ1B assay | 95% | Cizzle Bio (manufacturer) |
| Specificity of CIZ1B assay | 75% - 85% | Cizzle Bio (manufacturer) |
| Sensitivity of LDCT | 93% | (Dressler et al. 2024) |
| Specificity of LDCT | 84% | (Dressler et al. 2024) |

The model did not specifically account for adverse events (AE) from diagnostic procedures subsequent to LDCT as the Lokhandwala study incorporated some level of AE costs into their composite cost of diagnostic follow-up. Their study found that AEs could occur in 24% to 38% of cases, a factor that further contributes to barriers to LDCT adoption and both patient and clinician fear around the implications of a false positive result.

## Results

The results of the economic model demonstrate that the addition of the CIZ1B biomarker test as a secondary screening tool, either before or after LDCT in eligible, high-risk Medicare beneficiaries, could offer net savings in the range of $489M due to avoided unnecessary diagnostic procedures and the savings of identifying and treating cancers early. In a scenario where blood-based biomarker testing can also modestly expand the proportion of high-risk Medicare beneficiaries tested, from 4.6% to 5.3%, modeled savings grow to $518M. Both of these savings numbers factor in the cost of applying the CIZ1B test at a price of $295.

Figure 1 depicts the flow of the model given the available inputs and its impact in reducing unnecessary biopsies.

**Figure 1.**
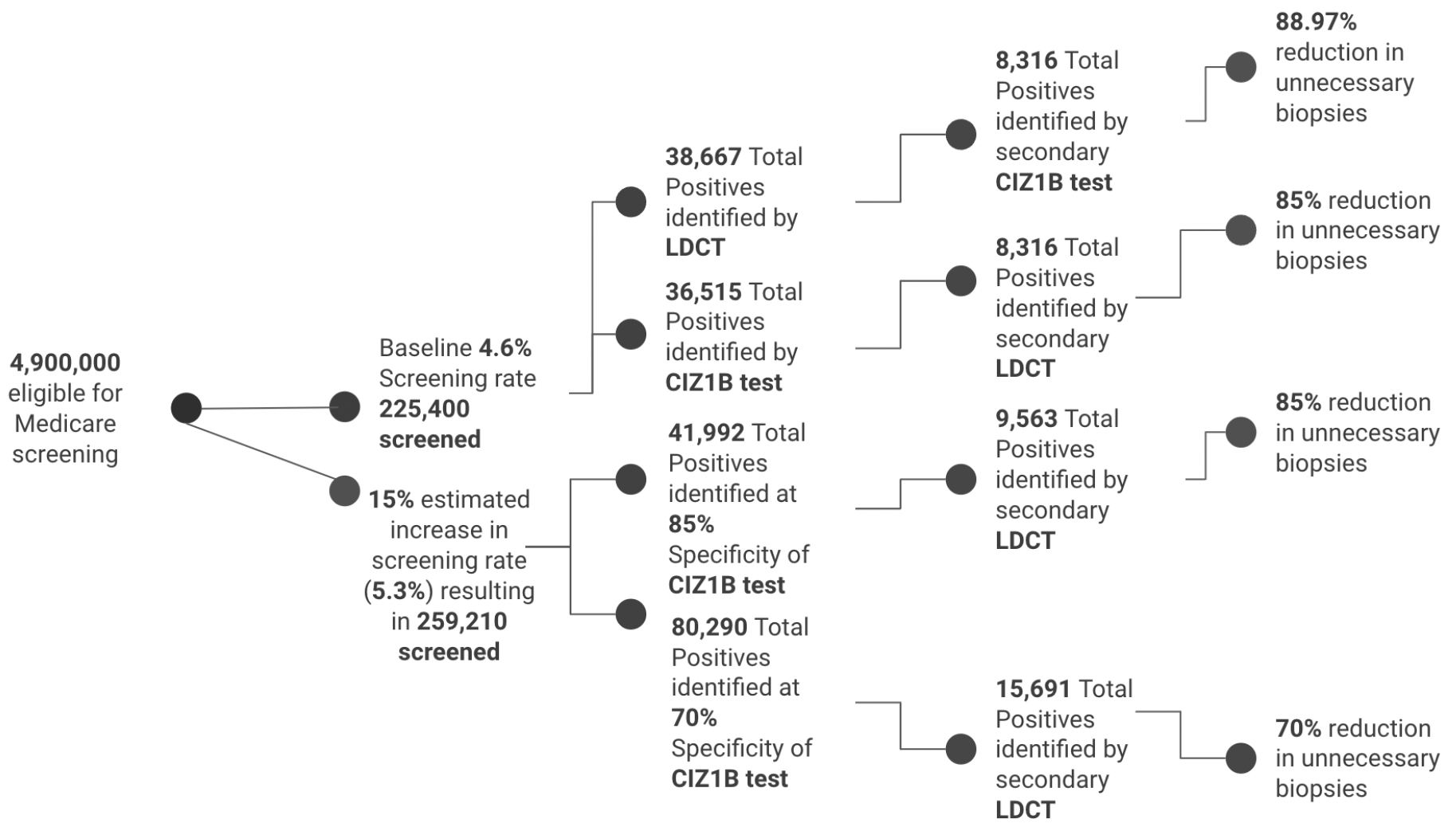

## Discussion

The persistently low uptake of LDCT screening represents a significant missed opportunity for early detection and intervention. In all cases, early detection not only offers clinical benefits, but also leads to lower costs as treatment of early-stage lung cancer is much less resource-intensive than treatment of late-stage LC. The reasons for low uptake are multifaceted, including both logistical barriers and other factors that deter participation in screening programs, as described earlier in this paper.

The introduction of a minimally invasive blood-based biomarker test could help overcome several of these barriers, including fear of unnecessary invasive procedures and logistical constraints. Any increase in early detection of LC cases at Stage I or II decreases cost burden and resource consumption, and more importantly, offers significant survival and mortality benefits for patients, as early stage LC is frequently curable.

Our modeling suggests that implementing CIZ1B testing could generate cost savings through two primary mechanisms:

1. Reduction in unnecessary procedures: The high rate of false positives in current LDCT screening leads to substantial costs for follow-up diagnostic procedures. More precise patient selection through adding a secondary screening mechanism biomarker testing could significantly reduce unnecessary invasive procedures.
2. Earlier detection: Detecting additional cases earlier (most likely at Stage I) rather than when symptomatic (often not until Stage IV) leads to significant cost savings as the treatment paradigm for early-stage LC is less resource-intensive and thus less costly, particularly as it does not involve high-cost systemic therapies.

The CIZ1B biomarker represents a promising advance in lung cancer screening. Previous research has demonstrated its capability to discriminate Stage I lung cancer from within high-risk groups with clinically useful accuracy, achieving receiver operator characteristic (ROC) areas under the curve (AUCs) exceeding 0.9 for two independent retrospective cohorts. (Higgins et al. 2012)

### Implementation Considerations

The success of implementing a CIZ1B biomarker test for lung cancer screening as complement to LDCT depends on several factors. To maximize adoption, cost to the Medicare beneficiary must be low to negligible, and ideally at $0 out of pocket to be on par with LDCT. Given the potential cost savings to Medicare of implementing this test, such a cost structure may be not only financially feasible but financially advantageous. Secondly, health care providers will need education about the role of biomarker screening in lung cancer and the clinical utility of a two-step screening paradigm, particularly to reduce false positives and subsequent invasive procedures. Lastly, blood-based testing could particularly benefit underserved populations who face barriers accessing LDCT screening facilities. The reduced logistical burden could help address current screening disparities and raise screening rates among many Medicare demographics.

### Model Limitations

Some limitations of our analysis should be noted:

1. Cost Data Uncertainty: Treatment costs continue to evolve rapidly with the introduction of new therapies, particularly immunotherapy combinations for advanced disease. Recent cost estimates for some components of our model were difficult to source from published data. Cost estimates may need regular updating to reflect current treatment patterns.
2. Implementation Assumptions: Our modeling assumes successful integration into clinical workflows. Real-world implementation challenges could affect the realized economic benefits, although these could be mitigated through payer policy and health care provider education as discussed.

### Considerations for Policy

USPTF guidance has historically moved slowly and been mired in bureaucracy as well as evidentiary debate; at the same time, oncology biomarker science is moving rapidly, evidentiary standards for clinical utility have been variable. Even cancer-focused groups such as National Comprehensive Cancer Network have struggled to keep up with the volume and speed of new oncology biomarker developments. Likewise, Medicare and private payers in the US have taken different approaches to coverage of biomarker testing, with significant policy action also occurring at the state level.

Any biomarker-based screening tool for lung cancer would likely require concerted effort on behalf of policy, clinician, and patient advocacy groups to gain wide adoption; understanding of significant clinical and economic benefit that might emerge from such tests could advance policy change in this regard. Economic benefits of early screening are positively compounded by increased screening rates and reduced barriers to screening access, particularly as LDCT rates in the US remain dismally low and frequently inequitable. Screening strategies that can both achieve better clinical outcomes in terms of reducing false positives and in potentially reducing barriers to screening, leading to positive health economic and public health results such as the CIZ1B biomarker test analyzed in this paper, offer several benefits attractive to policymakers.

## Data Availability

All data produced in the present study are available upon reasonable request to the authors

